# Orally administrated *Lactobacillus gasseri* TM13 and *Lactobacillus crispatus* LG55 Can Restore the Vaginal Health of Patients Recovering from Bacterial Vaginosis

**DOI:** 10.1101/2022.12.21.22283705

**Authors:** Fengyuan Qi, Shangrong Fan, Chao Fang, Lan Ge, Jinli Lyu, Zhuoqi Huang, Shaowei Zhao, Yuanqiang Zou, Liting Huang, Xinyang Liu, Yiheng Liang, Yongke Zhang, Yiyi Zhong, Haifeng Zhang, Liang Xiao, Xiaowei Zhang

## Abstract

Bacterial vaginosis (BV) is a common infection of the lower genital tract with a vaginal microbiome dysbiosis caused by decreasing of lactobacilli. Previous studies suggested that supplementation with live *Lactobacillus* may benefit the recovery of BV, while the outcomes vary in people from different regions. Herein, we aim to evaluate the effectiveness of oral Chinese-origin *Lactobacillus* with adjuvant metronidazole (MET) on treating Chinese BV patients. In total, 67 Chinese women with BV were enrolled in this parallel controlled trial and randomly assigned to two study groups: a control group treated with MET vaginal suppositories for 7 days and a probiotic group treated with oral *Lactobacillus gasseri* TM13 and *Lactobacillus crispatus* LG55 as an adjuvant to MET for 30 days. By comparing the participants with Nugent scores ≥ 7 and < 7 on days 14, 30, and 90, we found that oral administration of probiotics did not improve BV cure rates (57.14% and 67.74% at day 14, 57.14% and 58.06% at day 30, 32.14% and 48.39% at day 90 for probiotic and control group respectively). However, the probiotics were effective in restoring vaginal health after cure by showing higher proportion of participants with Nugent scores < 4 in the probiotic group compared to the control group (87.50% and 71.43% on day 14, 93.75% and 88.89% on day 30, and 77.78% and 66.67% on day 90). The relative abundance of the probiotic strains was significantly increased in the gut microbiome of the probiotic group compared to the control group at day 14, but no significance was detected after 30 and 90 days. Also, the probiotics were not detected in vaginal microbiome, suggesting that *L. gasseri* TM13 and *L. crispatus* LG55 mainly acted through the gut. A higher abundance of *Prevotella timonensis* at baseline was significantly associated with long-term cure failure of BV and greatly contributed to the enrichment of the lipid IVA synthesis pathway, which could aggravate inflammation response. To sum up, *L. gasseri* TM13 and *L. crispatus* LG55 can restore the vaginal health of patients recovering from BV, and individualized intervention mode should be developed to improve BV cure rates.

## 1 INTRODUCTION

Bacterial vaginosis (BV) is a microecological disorder caused by decreased abundance of lactobacilli and an increased abundance of anaerobic bacteria, commonly affecting the female lower genital tract (Ferreira et al., 2017). The prevalence of BV is around 20%-30% all over the world (Peebles et al., 2019). BV increases the susceptibility of women of reproductive age to sexually transmitted infections (STIs), including human immunodeficiency virus (HIV) (Smith et al., 2014), human papillomavirus (HPV) (Brotman et al., 2014), and gonorrhea (Bautista et al., 2016), and increases the risk of spontaneous abortion, preterm delivery, and amniotic fluid infection during pregnancy (Cherpes et al., 2003; Wiesenfeld et al., 2003; Donders et al., 2009; Cohen et al., 2012). Common symptoms of BV include elevated vaginal pH, increased leukorrhea, odor, vulvar itching, and burning pain. In addition, approximately 50% of patients are asymptomatic (Koumans et al., 2007; Ravel et al., 2013; Smith et al., 2014).

16S rRNA amplicon sequencing technology has been widely used to characterize the microbiome of the vagina in healthy or BV states (Brotman, 2011; Gajer et al., 2012; Vitali et al., 2015). In BV patients, *Lactobacillus* are replaced by anaerobic bacteria such as *Gardnerella spp*., *Prevotella spp*., *Mobiluncus spp*., and *Atopobium vaginae*, which result in high production of cadaveric amines, putrescine, succinate, and acetate leading to the altered chemical composition of secretions as well as lower pH (Xiao et al., 2016). However, 16S rRNA amplicon sequencing has low species resolution and functional information of microorganisms cannot be directly obtained from sequence data. Since metagenomic shotgun sequencing is able to identify microbial categories with high resolution and characterize their biological functions (Cao et al., 2020; Liu et al., 2020), it has been wildly used in explaining the structure and function of the vaginal microbiome (Liu et al., 2021; Ruiz-Perez et al., 2021). It’s no doubt that metagenomic shotgun sequencing is a better tool to study the mechanisms of vaginal microbiome involvement in pathogenesis of BV.

Metronidazole (MET) has been recommended by the treatment guidelines of BV (Workowski, 2015). Yet, the 12-month long-term cure rate is only 30% (Bostwick et al., 2016). The inhibition of MET penetration by biofilms is thought to be the main cause of BV recurrence (Togni et al., 2011). Probiotics have been used as an alternative treatment approach to prevent recurrent BV. For example, previous studies have reported that oral administration of *Lactobacillus* as an adjuvant therapy could increase the long-term cure rate of BV by improving the balance of the vaginal microbiome (Laue et al., 2018; Reznichenko et al., 2020; Chen et al., 2021; Martoni et al., 2022). Microbial translocation from the colon to the vagina has been hypothesized as a major pathway for the efficacy of oral probiotics (Reid et al., 2001; Morelli et al., 2004). Orally administrated probiotics could also suppress systemic inflammatory responses via the fermentation products of probiotics in the intestine (Amabebe and Anumba, 2020). However, the outcome of the probiotic treatment on BV could vary in different ethnic groups since the vaginal microbiome is different among women in different ethnic groups (Ravel et al., 2011). A study demonstrated that oral *L. rhamnosus* GR-1 and *L. reuteri* RC-14, isolated from European women, were barely detectable in the gut and vaginal microbiome after administration (Zhang et al., 2021). It has also been confirmed that native dominant *Lactobacillus* species could persistently colonize in the vagina compared to others (Vallor et al., 2001). This suggests that isolated probiotic strains are more likely to function in women of the same ethnic groups, but more evidences should be provided.

The probiotic strains used in this study are *L. gasseri* TM13 and *L. crispatus* LG55 which were isolated from the feces of 2 healthy Chinese people, which we developed in-house. They showed the strong ability to lower vaginal pH, inhibit the growth of pathogenic bacteria and fungi, and alleviated the inflammatory response of BV rats (Lyu et al., 2022). In this study, we aim at evaluating the effectiveness of oral Chinese-origin probiotic strains, *L. gasseri* TM13 and *L. crispatus* LG55, with adjuvant MET in treating Chinese BV patients, and investigate the dynamic of the gut and vaginal microbiome using metagenomic sequencing during the trial.

## 2 MATERIALS AND METHODS

### 2.1 Trial Population

Women who attended the gynecology outpatient clinic of Peking University Shenzhen Hospital in China between June 2020 and April 2021 and presented with abnormal leucorrhoea symptoms were enrolled in this single-center, prospective, parallel-group, randomized controlled clinical trial. Inclusion criteria were: age of 18 - 55 years and a history of sexual activity and a Nugent Score ≥ 7. Exclusion criteria were: vulvovaginal candidiasis (VVC), trichomonas vaginalis (TV) infection, *Chlamydia trachomatis* (CT) infection, gonococcal vaginitis, pregnant or planning to become pregnant, breastfeeding, pelvic inflammatory disease, allergic to MET, on antibiotic therapy, long-term contraceptive or immunosuppressive drug use or allergic, no regular sexual partner (RSP), and those with a history of systemic organic disease or psychiatric disorders.

This study was approved by the Peking University Shenzhen Hospital Medical Ethics Committee (ID: PUshenzhenH2020-009) and published on ClinicalTrials.gov (NCT04771728). Written consent was obtained from all subjects for this study.

### 2.2 Study Design

Patients with an initial Nugent Score ≥ 7 were informed of the study protocol. Those who met the criteria signed an informed consent form before the trial. Subjects were asked to complete a questionnaire containing information on demographic characteristics, vaginal health status, history of drug allergies, and history of reproductive system disorders. A random number table generated by SPSS 13.0 software was used to randomly assign subjects to the control or probiotic group using a 1:1 ratio. After enrollment, the probiotic group received MET vaginal suppositories (200 mg daily for 7 days) along with oral intake of probiotic solid drinks containing *L. gasseri* TM13 and *L. crispatus* LG55 (Lyu et al., 2022) (daily intake ≥ 5×10^9^ CFU for 30 days); the control group received the same dose of MET vaginal suppositories.

Follow-up visits were performed on day 14, 30, and 90 after the initiation of treatment. The investigator at enrolment distributed the intervention product. At each follow-up visit, the subject’s compliance, frequency of sex, vaginal health, and the occurrence of adverse events (AEs) were assessed by questionnaire. Subjects were required to avoid topical vaginal dosing during menstruation but continue to take the intervention product orally (**Figure 1A**). In addition, subjects were required to avoid sexual intercourse and vaginal douching throughout the follow-up period. We regrouped all subjects according to treatment outcomes. Subjects who were not found to be BV-negative at any of the follow-up visits were assigned to the treatment failure group; subjects who changed from BV-negative to BV-positive at any of the follow-up visits were assigned to the short-term cure group, and subjects who tested BV-negative at all follow-up visits were assigned to the long-term cure group.

**Figure 1.**
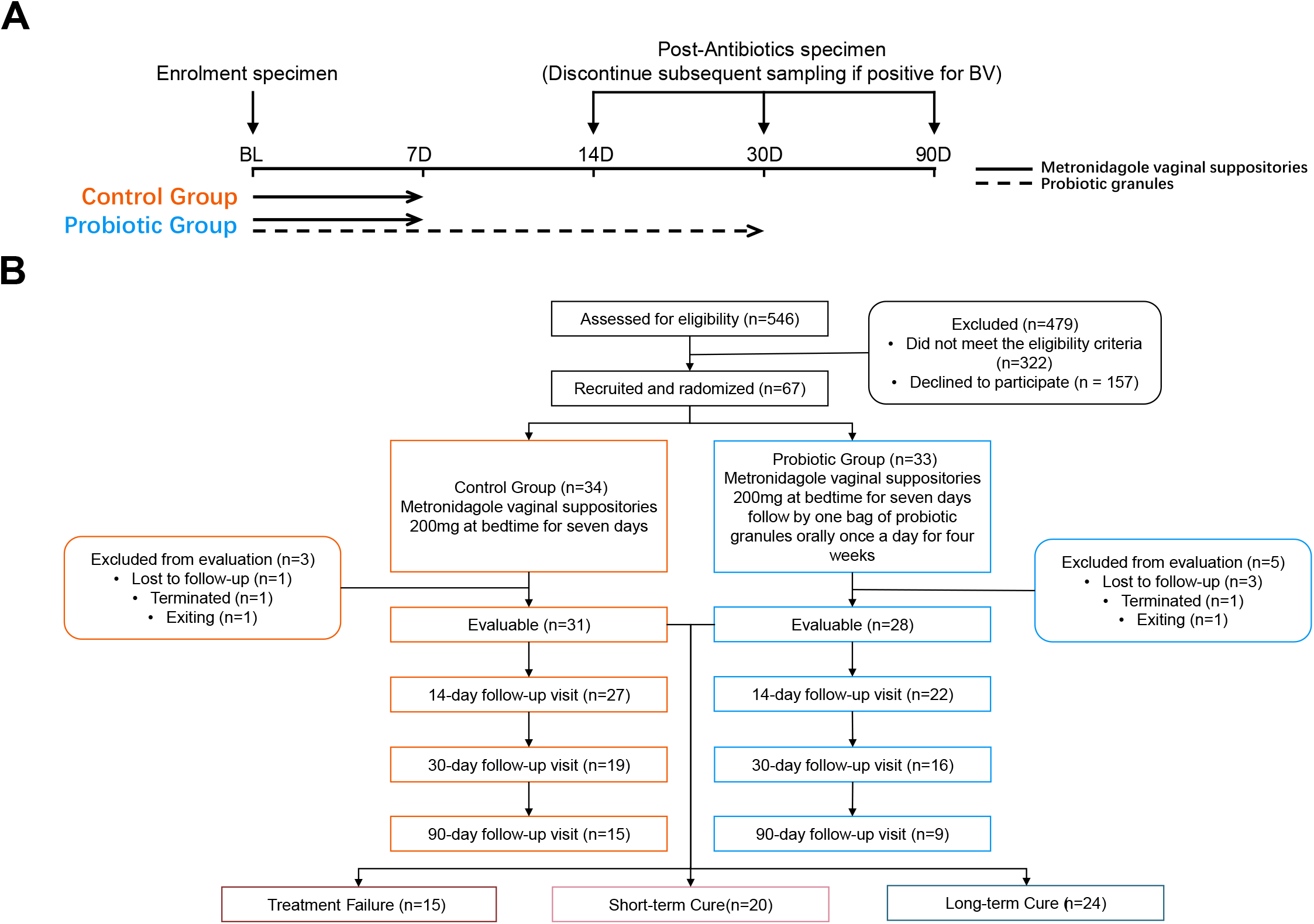
Study design **(A)** and participant flowchart **(B)**.

### 2.3 Efficacy Evaluation

Before treatment, all subjects were assessed for between-group differences in demographic characteristics, including age, height, weight, marital status, history of smoking, drug allergies, history of childbirth, and history of reproductive disorders. The diagnostic criteria for BV were based on the Nugent Score (Nugent et al., 1991) of vaginal smears. Gram-stained smears were examined under a microscope, and different morphological bacterial cells were counted at 1000× magnification: 0-6 points were diagnosed as BV negative and 7-10 points as BV positive. Two cytology technicians performed gram staining for a double-blind analysis. Clinical testing was also performed for pH, Donders LBG (Donders et al., 2002), and vaginal cleanliness grades. Efficacy outcomes were assessed by the percentage of subjects with no diagnosis of BV (diagnostic criteria of Nugent Score ≤ 7) at each follow-up visit as the cure rate, based on compliance with protocol set (PP) analysis. Those with a Nugent Score ≥7 were excluded from the clinical trial at each follow-up visit, but these subjects were still included when we assessed cure rates at subsequent follow-up time points.

### 2.4 Metagenomic Sequencing and Biological Information Annotation

Fecal and vaginal samples collected from participants during the initial and follow-up consultations were subjected to DNA extraction and metagenomic shotgun sequencing. Metagenomic shotgun sequencing (100bp paired reads) was performed on the DNBSEQ™ platform (Fang et al., 2018; Han et al., 2018; Chen et al., 2021). Sequencing data were quality-controlled using fastp v0.20.1 (http://github.com/OpenGene/fastp) (Chen et al., 2018), filtering low-quality sequences with default parameters. High-quality sequences were then aligned to the hg38 human reference gene set using Bowtie2 v2.4.2 (http://github.com/BenLangmead/bowtie2) (Langmead et al., 2009) to exclude the host genome. The de-hosted high-quality sequences were aligned using MetaPhlAn v3.0.7 (http://huttenhower.sph.harvard.edu/metaphlan) as well as HUMAnN 3.0 (http://huttenhower.sph.harvard.edu/humann3) for taxonomic as well as functional annotation (Beghini et al., 2021).

### 2.5 Statistical Analysis

The Shannon and Simpson index and Bray-Curtis distance were used to calculate alpha diversity and beta diversity based on the relative abundance of species. The t-test and Mann-Whitney U-test were used to test for differences between two numerical variables in two groups, with 0.05 considered as the threshold for significant differences in p-values. The Chi-square test and Fisher’s exact test were used for differences between groups for categorical variables. Ward Linkage hierarchical clustering of species-level relative abundance of vaginal microorganisms using R Stats v3.5.3 with Jensen-Shannon distances. LEfSe (Segata et al., 2011) was used to identify species that differed between different treatment outcomes, and a general linear model by MaAsLin2 (Mallick et al., 2021) was used to look for the significantly different pathways. In order to construct co-occurrence networks, we first screened for species with mean relative abundance ≥ 0.1%, then calculated interspecific Spearman correlation coefficients, and constructed co-occurrence networks for two different treatment outcome groups using all correlations with p-values ≤ 0.05, which were visualized by Gephi.

## 3 RESULTS

### 3.1 Clinical Trial Process and Efficacy Evaluation

After screening 546 BV patients with Nugent Score ≥7 according to the inclusion and exclusion criteria, 67 patients were included in the clinical trial and randomized. Among them, 8 subjects were excluded (4 subjects were lost to follow-up, 2 subjects were terminated due to non-medication as prescribed, and 2 subjects withdrew from this study for personal reasons). Ultimately, 31 subjects in the control group and 28 subjects in the probiotic group were included in the efficacy evaluation and microbiome analysis, along with 31 subjects in the control group (**Figure 1B**). Subjects’ demographic and pathological characteristics at baseline did not differ significantly between the probiotic and control groups (all P > 0.05) (**Table 1**).

**Table 1.** Demographic and behavioral characteristics of subjects at baseline.

|  | Probiotic group<br>(n=28) | Control group<br>(n=31) | *P* |
| --- | --- | --- | --- |
| individual characteristic |  |  |  |
| Age (years, mean ± SD) | 34.3 ± 7.7 | 32.1 ± 6.8 | 0.249 |
| Height (cm, mean ± SD) | 160.9 ± 5.8 | 161 ± 4.9 | 0.943 |
| Weight (kg, mean ± SD) | 54.7 ± 14.0 | 51.7 ± 8.6 | 0.320 |
| Smoking history, n (%) | 1 (3.57) | 3 (9.68) | 0.614 |
| Current relationship status, n (%) |  |  |  |
| Married | 19 (67.86) | 16 (51.61) | 0.289 |
| Single, divorced or others | 9 (32.14) | 15 (48.39) |  |
| Contraception, n (%) | 21 (75.00) | 15 (48.39) | 0.060 |
| Pregnancies num (n, mean ± SD) | 2.2 ± 1.8 | 1.6 ± 1.7 | 0.130 |
| Delivery num | 1.1 ± 1.0 | 0.8 ± 1.1 | 0.279 |
| Clinical symptoms or signs, n (%) |  |  |  |
| leucorrhea | 26 (92.85) | 31 (100) | 0.221 |
| Fishy smell or other bad Vaginal odour | 27 (96.43) | 31 (100) | 0.475 |
| Itching | 8 (28.57) | 7 (22.58) | 0.564 |
| Burning | 1 (3.57) | 1 (3.23) | 1.000 |
| Laboratory test |  |  |  |
| Cleaning degree, n (%) |  |  |  |
| 3 | 27 (96.4) | 29 (93.5) | 1.000 |
| 4 | 1 (3.57) | 2 (6.45) |  |
| Nugent score (mean ± SD) | 8.04 ± 0.64 | 8.03 ± 0.71 | 0.955 |
| Donders , n (%) |  |  |  |
| I | 1 (3.57) | 0 (0) | 0.567 |
| II b | 1 (3.57) | 1 (3.23) |  |
| III | 26 (92.85) | 30 (96.77) |  |
| Vaginal pH (mean ± SD) | 4.78 ± 0.16 | 4.86 ± 0.22 | 0.119 |

At all time-points, there was no significant improvement in the cure rate in the probiotic group compared to the control group. The cure rates for the probiotic and control groups were 72.73% and 84.00% at day 14, respectively; 57.14%, and 60.00% at day 30, respectively; 32.14% and 48.39% at day 90, respectively. Notably, in the group of cured participants, the proportion of those fully recovered (NS < 4) was higher in the probiotic group compared to the control group (87.50% and 71.43% on day 14, 93.75% and 88.89% on day 30, and 77.78% and 66.67% on day 90, respectively) (**Figure 2A**). To sum up, this suggests that oral administration of *L. gasseri* TM13 with *L. crispatus* LG55 cannot improve BV cure rates but has a role in restoring vaginal health after cure. We also detected BV-associated symptoms and laboratory parameters at different time-point of the trial, however, no significant inter-group differences were found (**Figure 2B**).

**Figure 2.**
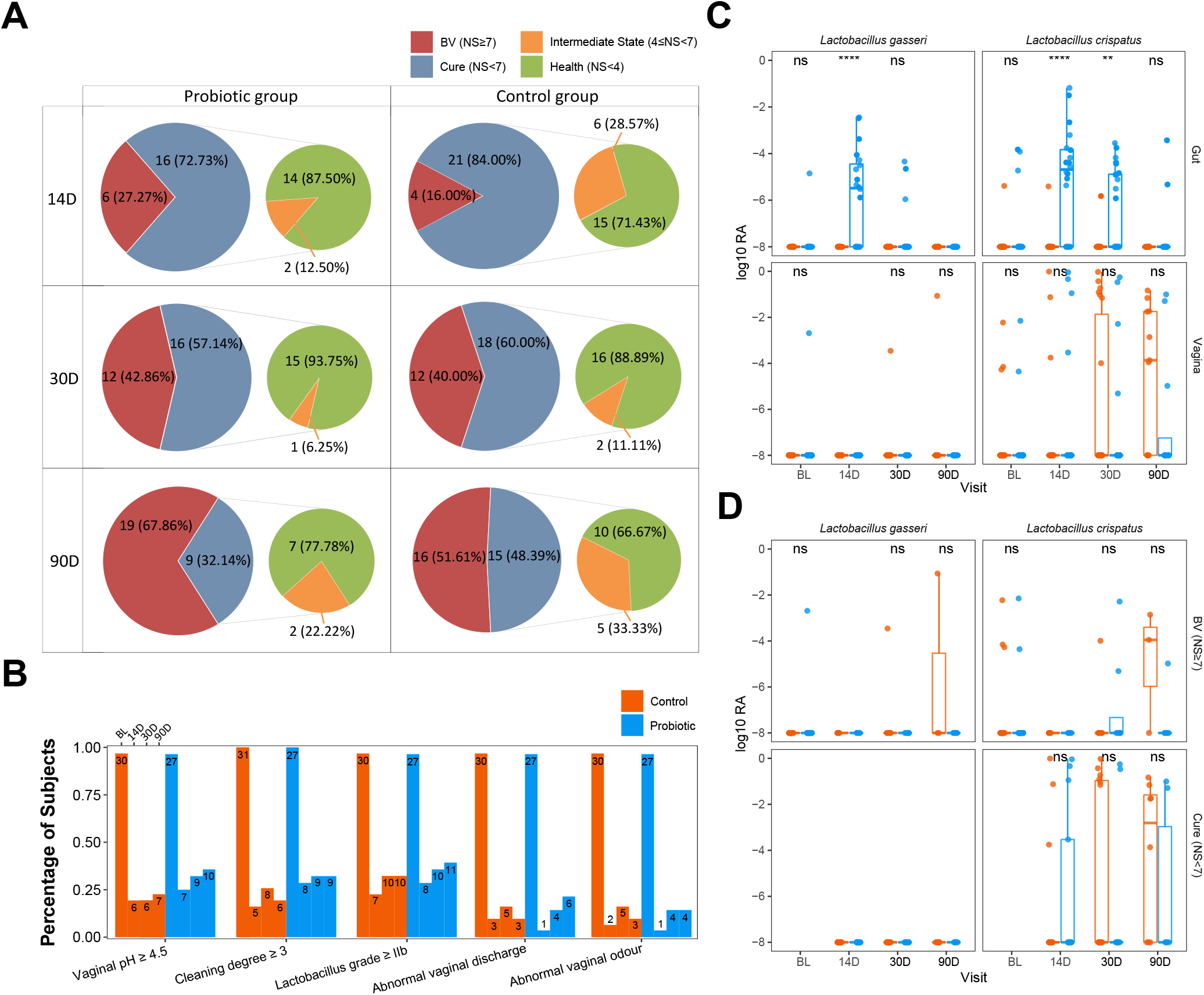
Although orally administrated *L. gasseri* TM13 and *L. crispatus* LG55 cannot improve BV cure rates, it restores the vaginal health after cure mainly acting through the gut. The number and percentage of the participants with different disease states (BV, non-BV, health, and transition stage) at different time points of the trial were shown in pie charts **(A)**. The percentage of the participants with detected BV-associated symptoms (abnormal vaginal discharge and abnormal vaginal odour) and laboratory parameters (vaginal pH, vaginal cleaning degree and Donders *Lactobacillus* grade) at different time-point of the trial were shown as a histogram **(B)**. The number marked on the top of the bar denote the case number. Box-and-whisker plots showing the relative abundance of intervention species between groups at different time-points in the gut or vagina **(C)**. All vaginal samples were grouped according to the Nugent scores ≥ 7 (BV) and < 7 (cure) **(D)**. Mann-Whitney U test were used to perform the statistical analysis. * stands for P < 0.05; ** stands for P < 0.01; *** stands for P < 0.001.

### 3.2 Inter-group Differences in the Abundance and Microbial Diversity of the Intervention Strains

In the fecal samples, the relative abundance of *L. crispatus* and *L. gasseri* was significantly greater in the probiotic group than in the control group on day 14 (P < 0.001). On day 30, only *L. crispatus* remained significantly different (P = 0.0037), and the relative abundance of both species in the probiotic group decreased compared to day 14. On day 90, the presence of the *L. gasseri* was not observed in the gut, and the presence of the *L. crispatus* was detected in only 2 participants (**Figure 2C**). This indicated that the colonization ability of *L. gasseri* TM13 in gut was not as strong as *L. crispatus* LG55. Furthermore, there was no difference in the relative abundance of *L. crispatus* and *L. gasseri* in the vaginal microbiome between probiotic and control groups (all P > 0.05) (**Figure 2C**), as well as between BV and cure groups (**Figure 2D**), which suggested that no transfer of the intervention strains from the gut to the vagina were detected.

In addition, to investigate the effect of oral probiotics on the microbial community structure of the gut and vagina, we observed the differences in alpha-diversity and beta-diversity in the gut and vaginal microbiome before and after the intervention. No difference in microbial diversity was found between the probiotic and control groups, either in the gut or the vaginal microbiome (all P > 0.05) (**Figure S1**).

### 3.3 Heterogeneity of the vaginal microbiome and association with treatment outcomes

All of the vaginal samples were grouped into five clusters by hierarchical clustering method (**Figure 3A**). *Lactobacillus iners* was dominant in Cluster 1 (37.16%), and the dominant species in Cluster 4 (6.76%) was vary in samples, including *L. crispatus, L. jensenii* and other species with lower average relative abundance. Cluster 2, Cluster 3, and Cluster 5 were highly associated with BV-associated pathogenic bacteria, including *Gardnerella, Prevotella*, and *Atopobium* dominated. Among them, a high abundance of *Prevotella amnii* was the main feature of Cluster 3 (12.84%), and Cluster 2 (24.32%) was dominated by *Gardnerella vaginalis*. While *Prevotella bivia, Prevotella timonensis*, and *Atopobium vaginae* were the main components of BV-associated pathogens in Cluster 5 (18.92%) and there was no clearly dominant species. It is noteworthy that baseline samples presented more frequently in Cluster 3 (80%) than in Cluster 2 (50.00%) or Cluster 3 (50.00%) (P = 0.02&0.03), which demonstrated that distribution regularity could exist in the baseline microbiome (BV microbiome) that may influence the treatment outcomes. To further investigate this issue, we selected the vaginal samples at baseline and re-clustered them according to the outcomes (**Figure 3B**). The result showed that the Cluster composition differed nearly significantly between long-term cure group and short-term cure group (P = 0.059). This suggested a strong relationship between the BV microbiome and long-term cure rate of BV, which could result in the treatment outcomes of *L. gasseri* TM13 and *L. crispatus* LG55.

**Figure 3.**
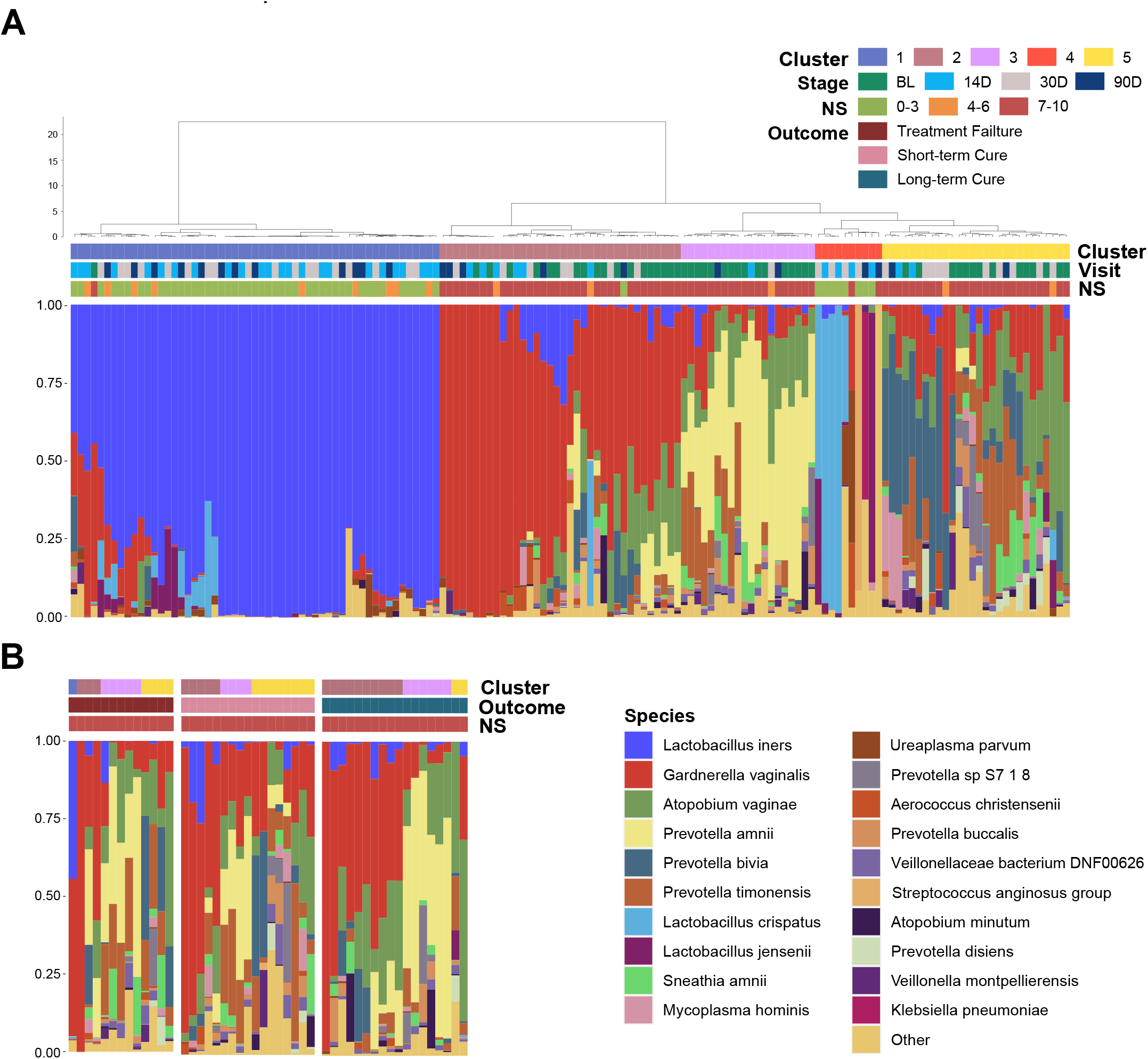
Distribution of the baseline microbiome (BV microbiome) greatly associated with the treatment outcomes. The vaginal microbiome of 59 women sampled longitudinally at all time points **(A)** and at baseline **(B)**. The samples are hierarchically clustered (R base hclust function with Jensen-Shannon distances and Ward linkage) and classified into five clusters. Nugent score range, follow-up time-points and treatment outcomes were indicated by the bars.

### 3.4 Differences in Vaginal Microbiome between the Short-term Cured Subjects and the Long-term Cured Subjects

To describe the differences in core species distribution and their interactions between the short-term cure group (**Figure 4A**) and the long-term cure group (**Figure 4B**), the high-abundance species (mean relative abundance ≥0.1%) in these two groups were selected to perform the co-occurrence network analysis. A greater number of high-abundance species of BV-associated pathogenic bacteria and a greater density of positive interspecies interactions was shown in the short-term cure group (106 significant positive correlations) than in long-term cure group (53 significant positive correlations). This suggests that stronger interbacterial interactions help them escape the antibiotic, which is in line with the findings of Gustin’s study (Gustin et al., 2022). *Lactobacillus*, which possessed higher abundance in the cured samples, had the most negative correlations due to the ability of *Lactobacillus* to secrete lactic acid and bactericidal substances, which inhibit the growth of BV-associated pathogenic bacteria (Zhu et al., 2022). In addition, we found *Ureaplasma parvum* enriched in cured samples both in the short-term cured group and in the long-term cured group and a significant negative correlation with certain BV-associated bacteria, which is consistent with the study of Xiao *et al*. (Xiao et al., 2019). With its small genome (Paralanov et al., 2012) and lack of biosynthetic capacity (Combaz-Sohnchen and Kuhn, 2017), we speculate that *Ureaplasma parvum* inhibits the growth of BV-associated pathogenic bacteria by activating the host immune response.

**Figure 4.**
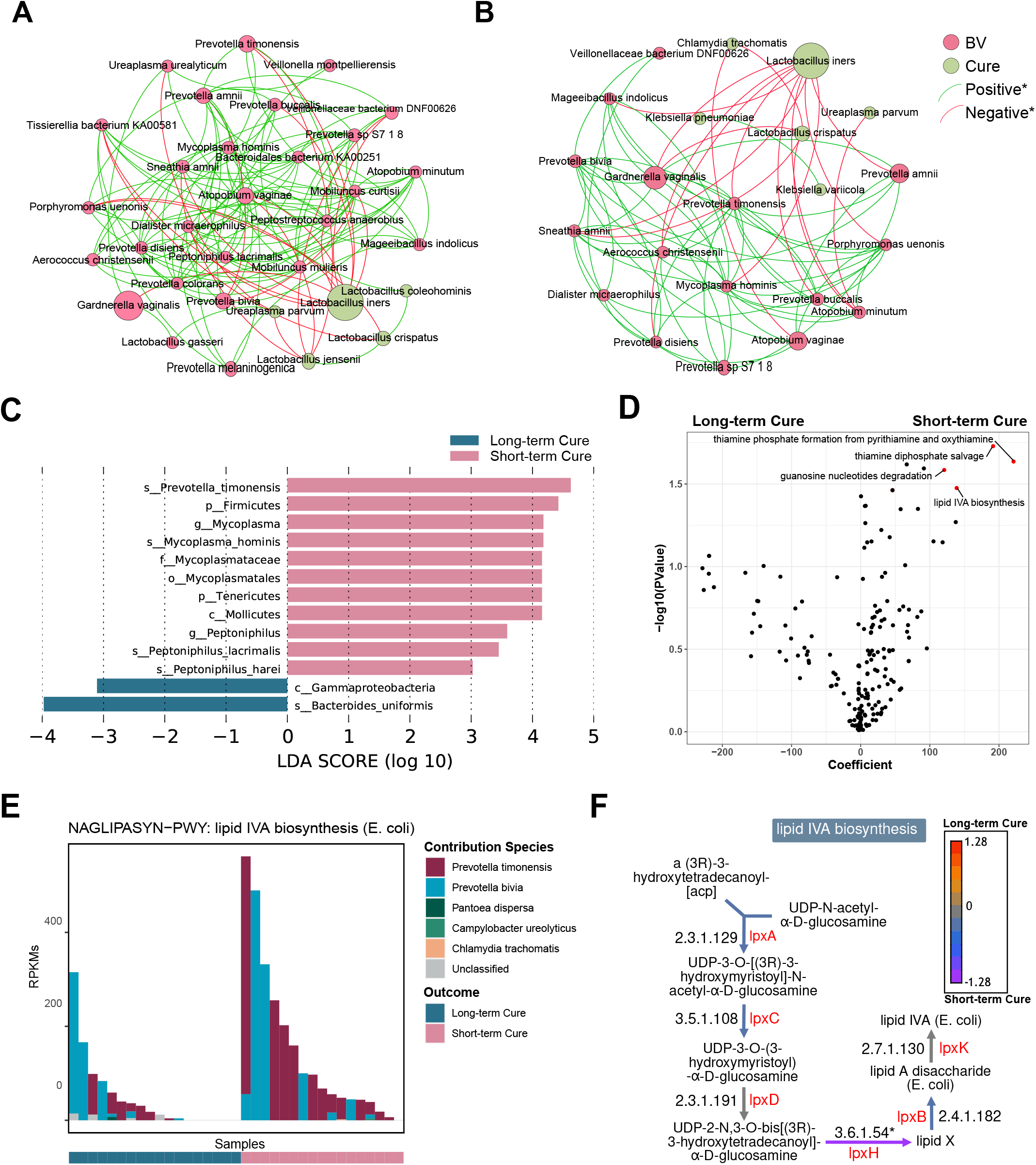
A higher abundance of *Prevotella timonensis* at baseline was significantly associated with long-term cure failure of BV and greatly contributed to the enrichment of the lipid IVA synthesis pathway. Co-occurrence network showing the interbacterial correlations of vaginal microbiome in the short-term cure group **(A)** and long-term cure group **(B)**. Each species is only shown in a color corresponding to the time-point when it has the highest relative abundance. Node size indicates the average abundance of each genus. Lines between nodes represent the interbacterial correlations, and the green line and red line indicate positive and negative correlations, respectively. Histogram of the linear discriminant analysis (LDA) scores computed for species **(C)** with differential abundance between the short-term cure group and long-term group in baseline, and the differential pathways **(D)** found by the linear model (points with p < 0.05 were marked in red). The contribution ranking of the vaginal microbiome to the lipid IVA biosynthesis pathway is shown in the stack bar chart **(E)**. The top of each set of stacked bars indicates the total abundance of the pathway within a single sample. The individual enzymatic steps of the lipid IVA biosynthesis pathway, as well as the differences between the short-term cure group and long-term group of the gene expressing these enzymes, are shown in this flowchart **(F)**. Mann-Whitney U test; *stands for p-value < 0.05; **stands for p-value < 0.01; ***stands for p-value < 0.001.

LEfSe analysis showed differences in vaginal microbiome at baseline between the short-term and long-term cure groups (**Figure 4C**). LDA scores indicated that *Prevotella timonensis, Mycoplasma hominis, Peptoniphilus lacrimalis*, and *Peptoniphilus harei* were more abundant in the short-term cure; *Bacteroides uniformis* was more abundant in the group with long-term cure. Using the generalized linear model in the MaAsLin2 package, we performed an association analysis of gene copy numbers of functional pathways in the vaginal microbiome at baseline (**Figure 4D**). In the short-term cure group, the gene copy number of lipid IVA synthesis, thiamine phosphate synthesis, and thiamine diphosphate salvage were more abundant than those in the long-term cure group (P < 0.05). Among them, the enrichment of the lipid IVA synthesis pathway in the short-term cure group was mainly caused by the high expression of UDP-2,3-diacylglucosamine diphosphatase encoded by lpxH (**Figure 4F**), and the greatest species contribution was mainly provided by *Prevotella timonensis* **(Figure 4E)**, which was consistent with the results of LEfSe analysis.

## 4 DISCUSSION

In this study, there was no significant difference in BV cure rates between the probiotic and control groups at day 14, day 30, and day 90, suggesting a 30-day oral administration of *L. gasseri* TM13 and *L. crispatus* LG55 was ineffective as an adjuvant treatment of BV. The short intervention period may be one of the main reasons for the ineffectiveness compared to other studies that obtained effective intervention results (Cohen et al., 2020; Reznichenko et al., 2020). In addition, in the BV conversion population, the percentage of people in a transition state was lower in the probiotic group at all three time points. This suggests that oral probiotics can modulate and improve vaginal health in cured subjects, which is consistent with previous studies (Chen et al., 2021; Martoni et al., 2022). Hence, oral administration of *L. gasseri* TM13 and *L. crispatus* LG55 is effective in restoring the vaginal health of patients recovered from BV.

The inability of the orally administered probiotic strains to reach or colonized to vagina may be the reason of the treatment failure in BV. Similarly, Husain *et al*. showed that the colonization rate of the intervention strains and the diversity of the vaginal microbiome did not differ between the probiotic and control groups (Husain et al., 2020). In fact, *L. gasseri TM13* and *L. crispatus LG55* used in this study were isolated, cultured, and screened from the human intestine (Lyu et al., 2022). Further evaluation on whether the differences in the physicochemical environment and differences in patterns of interactions between bacteria in the vagina and the intestine (Human Microbiome Project, 2012) can prevent the transfer of the intervening strains from the intestine to the vagina is needed. Previous studies have detected probiotics in the vagina using qPCR (Cohen et al., 2020) and 16s rRNA amplicon sequencing (Martoni et al., 2022), but these methods cannot provide strain-specific abundance information, so the conclusions are unreliable. In recent years, strain identification methods based on metagenome sequencing data have emerged, such as MetaMp (Dilthey et al., 2019), which uses long-read sequencing technology for strain-level annotation, PStrain (Wang et al., 2020), based on single nucleotide variants (SNVs), and StrainPanDA (Hu et al., 2022) that uses pangenome. These advanced methods can provide a more accurate quantitative detection of probiotic colonization and can give direct evidence of the mechanism of action of orally administered probiotics *in vivo*.

In this study, there was no difference in the relative abundance of *L. crispatus* and *L. gasseri* in the vaginal microbiome between probiotic and control groups at all follow-up time point, which suggest that *L. gasseri* TM13 and *L. crispatus* LG55 mainly influenced the host immune response through the gut. Oral administration of *L. gasseri* CECT5714 increases the production of SCFAs in the intestine (Olivares et al., 2006) and SCFAs exert anti-inflammatory effects by modulating the levels of PGE, cytokines, and chemokines (Cox et al., 2009). In addition, consuming fermented milk containing *L. crispatus SMFM2016-NK* can effectively reduce the expression levels of intestinal TNFα and IL-1β (Choi et al., 2021). Therefore, *L. gasseri* TM13 and *L. crispatus* LG55 may also exert their effects on the improvement of vaginal health by reducing systemic inflammation through immunomodulatory effects in the intestine. In the present study, the abundance of the intervening strains in the intestine could not be maintained at high levels even during the 30-day intervention period. This might be related to the endogenous stability and resilience of the intestinal flora (Lozupone et al., 2012), which can prevent the long-term colonization of the intervening strains in the intestine, thus preventing this bacteria from exerting probiotic functions in a sustained and stable manner. Our results also support this inference; the largest difference between the probiotic and control groups in the proportion of participants with NS<4 occurred on day 14. Therefore, to improve the effect of *L. gasseri TM13* and *L. crispatus LG55*, a longer intervention period might be one of the optimization options.

The abundance of key bacteria in the vagina influences whether BV can be cured in the long term (Xiao et al., 2019; Mollin et al., 2022). We found that *Prevotella timonensis* is enriched in the vagina at the baseline in long-term cure failure patients and that the lipid IVA synthesis pathway, which is abundantly enriched in *Prevotella timonensis*, is an important precursor material for LPS biosynthesis. *Prevotella timonensis* is a strictly anaerobic Gram-negative bacterium enriched in the genital tract of BV (Lehtoranta et al., 2020), Chlamydia trachomatis infection (Filardo et al., 2019), and HPV infection (Chen et al., 2020).Nienke *et al*. showed that *Prevotella timonensis* induces the maturation of DC cells to secrete large amounts of pro-inflammatory cytokines, including IL-1β and IL-8, to enhance the inflammatory response in the genital tract (van Teijlingen et al., 2020). Our results also showed a significantly positive correlation between *Prevotella timonensis* and Lipid IVA synthesis. Lipid IVA is the lipopolysaccharide (LPS) A tetra-acetylated precursor of lipid A in biosynthesis. Although Lipid IVA has been reported as a structural antagonist of LPS, it could also be a substrate of LPS synthesis and further activated the innate immune responses via TLR4 and its co-receptor MD-2 (Ohto et al., 2012). We speculate that *Prevotella timonensis* aggravated the inflammatory effect by producing LPS persistently, which would induce long-term cure failure of BV. But the pro-inflammatory mechanism of *Prevotella timonensis* in the vagina and how it perturbed the vaginal microbiome and host immune response still need further study. Furthermore, the probiotic effect of oral *L. gasseri TM13* and *L. crispatus LG55* on the genital tract has a limited effect in curing subjects, suggesting that precise treatment and personalized probiotic interventions are necessary for BV patients.

This study has some limitations. In terms of the clinical trial design, the participants completed their trials when diagnosed with BV, and there was no continuous follow-up for these patients. We were unable to provide a comprehensive description of the microbiome dynamics of BV treatment failure. Regarding bioinformatics analysis techniques, the relative abundance at the species level was insufficient to accurately calculate the number of intervention probiotic strains in the samples. In addition, the relatively small sample size is also one of the limitations of this study. Finally, future microbiome studies on probiotic interventions to assist in treating BV need to be supported by larger sample sizes and strain-level annotation techniques with higher resolution.

## 5 CONCLUSIONS

Although orally administrated *L. gasseri* TM13 and *L. crispatus* LG55 cannot improve BV cure rates, it restores vaginal health after cure mainly acting through the gut. A higher abundance of *Prevotella timonensis* at baseline was significantly associated with long-term cure failure of BV and greatly contributed to the enrichment of the lipid IVA synthesis pathway, which could aggravate inflammation response. This inferred that individualized intervention mode should be developed to improve BV cure rates.

## Data Availability

The data that support the findings of this study have been deposited into the CNGB Sequence Archive of China National GeneBank DataBase with accession number CNP0003852.

## 6 DATA AVAILABILITY STATEMENT

The data that support the findings of this study have been deposited into the CNGB Sequence Archive of China National GeneBank DataBase with accession number CNP0003852 (https://db.cngb.org/search/project/CNP0003852/).

## 7 ETHICS STATEMENT

The studies involving human participants were reviewed and approved by the Medical Ethnic Committee of Peking University Shenzhen Hospital (ID: PUshenzhenH2020-009). The patients/participants provided their written informed consent to participate in this study.

## 9 AUTHOR CONTRIBUTIONS

XZ, LX and LG designed the research. SF, JL, LH, XL, YL and YZ carried out the clinical trial. XZ, LG, FQ, JL and SZ supported the clinical trial and organized data. FQ, CF, ZH and SZ analyzed the data. FQ, XZ, CF and ZH draft the paper. LX, SF and LG provided critical revisions of the article. LG, YZ and HZ were responsible for the probiotic product support. XZ and XL supervised the project. All authors contributed to the article and approved the submitted version.

## 11 ACKNOWLEDGMENT

We thank the principal investigators, researchers, and all the participants. This work was supported by China National GeneBank.

## SUPPLEMENTARY MATERIAL

**Figure S1:** Shannon and Simpson index between the probiotics group and the control group in all time points **(A)**. PCoA-based Bray-Curtis distance matrices between the probiotics group and the control group in all time points **(B)**.

