## Supplemental Figure 1 for "Orally administrated *Lactobacillus gasseri* TM13 and *Lactobacillus crispatus* LG55 Can Restore the Vaginal Health of Patients Recovering from Bacterial Vaginosis"

**Figure S1:** Shannon and Simpson index between the probiotics group and the control group in all time points **(A)**. PCoA-based Bray-Curtis distance matrices between the probiotics group and the control group in all time points **(B)**.

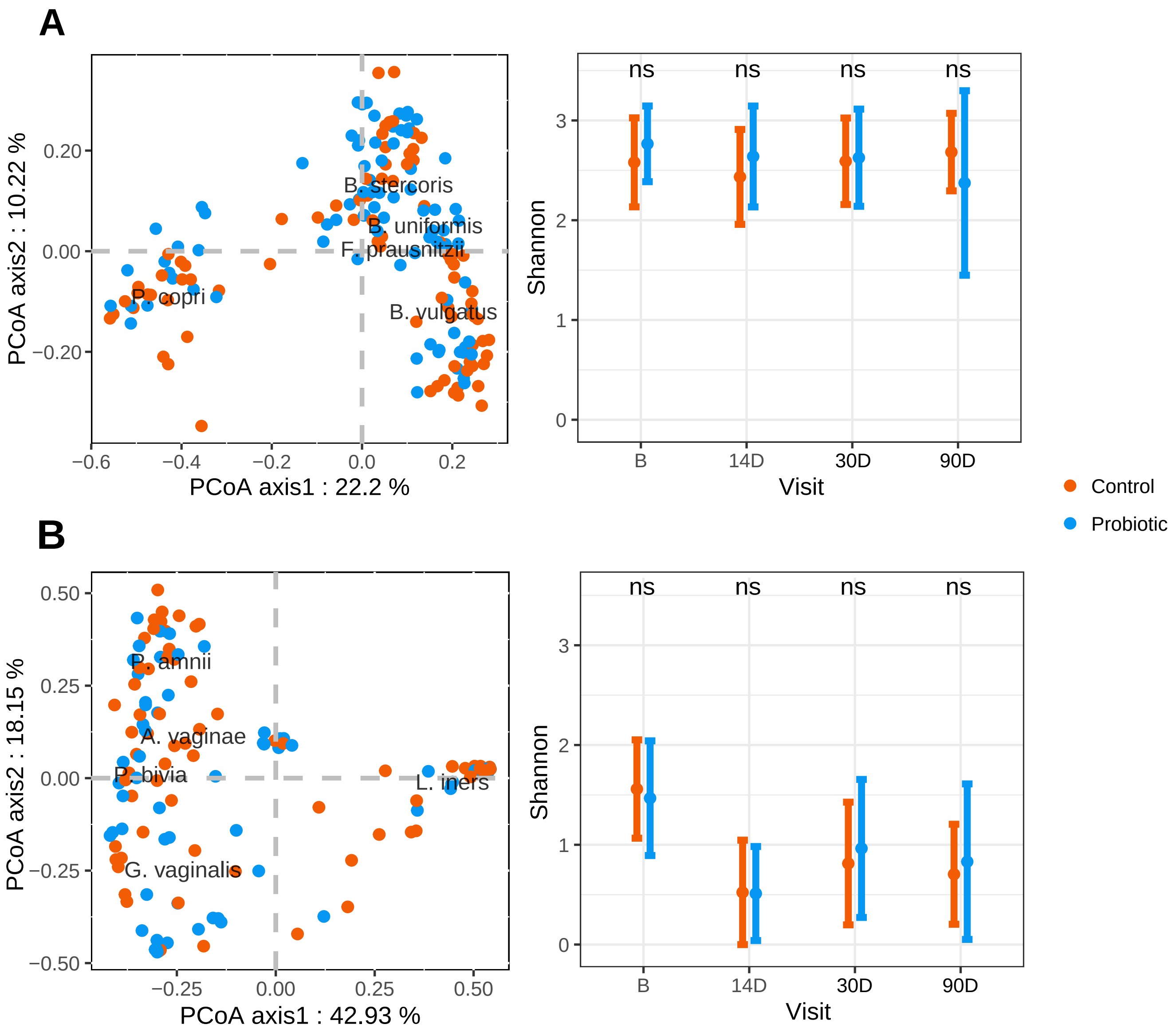
